# Implementation of a national oxygen distribution network in Lesotho: a longitudinal analysis

**DOI:** 10.1101/2024.09.05.24313130

**Authors:** Melino Ndayizigiye, Afom T Andom, Palesa Thabane, Mphatso Tsoka, Francis Sambani, Tumelo Monyane, Juliana Lawrence, Ninza Sheyo, Mpho Pholoanyane, Jessica Parker, William Haggerty, Emily Gingras, Tiara Calhoun, Joia Mukherjee, Paul D Sonenthal

## Abstract

**Background:** Despite its essential and life-saving role in the treatment of many medical conditions, access to medical oxygen remains limited in many countries. In 2021, Partners In Health established an oxygen distribution network in Lesotho to increase medical oxygen access.

**Methods:** We conducted a longitudinal analysis of the implementation of an oxygen distribution network in Lesotho from November 2022 to January 2024. Oxygen delivery data were abstracted from tracking logs and analysed in Stata. Continuous and ordinal variables were summarized by medians and ranges. Categorical variables were described using frequencies and proportions.

**Results:** Over the 15-month study period, the network expanded from 1 oxygen production hub serving 5 recipients to 4 hubs and 21 recipients located across nine of Lesotho’s ten districts. The network delivered 1,565 filled cylinders containing 9,619.23 m^3^ oxygen. For the 13 recipients with inpatient beds, the median monthly volume of oxygen delivered per bed was 1.43 m^3^ (IQR: 0.57 to 2.31).

**Conclusion:** This study demonstrates the feasibility and impact of an oxygen distribution network in Lesotho, providing proof-of-concept for an intervention to improve oxygen access in LMICs. By employing real-time monitoring and redundant sourcing, the network provided a reliable oxygen supply responsive to variations in demand and periods of oxygen plant downtime. This study also provides insights into facility-level oxygen consumption, which may help policymakers improve quantification and prediction of oxygen demand. Future efforts should focus on enhancing data collection, characterizing oxygen usage, and strengthening infrastructure to promote sustainable oxygen security.

**KEY QUESTIONS:** *What is already known on this topic:* Improving access to medical oxygen is a global health priority, as underscored by World Health Assembly Resolution 76.3. Existing research has largely concentrated on facility-level interventions, establishing the impact of multifaceted strategies such as distributing oxygen concentrators, training clinicians, and strengthening supply chains.

*What this study adds:* To our knowledge, this is the first study to provide detailed prospective data on the operation of a national oxygen distribution system utilizing a novel “network” model integrating multiple production hubs. It demonstrates the feasibility and effectiveness of this approach, showing improved resilience and reliability in medical oxygen supply across a broad geographic area in a LMIC.

*How this study might affect research, practice or policy:* Our results demonstrate that oxygen distribution networks with multiple hubs can help meet regional and national oxygen demands, providing a resilient solution for strengthening oxygen ecosystems by dynamically aligning supply and demand. Additionally, these networks facilitate enhanced data collection for estimating oxygen demand, thereby informing future policy and implementation strategies.

## INTRODUCTION

Medical oxygen, recognized by the World Health Organization (WHO) as an essential medication,[1] is critical for treatment of hypoxemia, a complication of numerous diseases, from neonatal respiratory distress to tuberculosis to chronic obstructive pulmonary disease. However, many healthcare facilities, particularly in low- and middle-income countries (LMICs), lack adequate access to medical oxygen, leading to preventable morbidity, often disproportionately affecting vulnerable populations such as children.[2–5] Developing innovative approaches to oxygen delivery has been identified as an intervention that can save lives[5,6] and help achieve Universal Health Coverage.[3]

The COVID-19 pandemic highlighted and exacerbated existing disparities in oxygen access, with many LMICs struggling to meet the soaring demand for medical oxygen. A study of 3,140 adults with suspected or confirmed COVID-19 infection admitted to intensive or high-care units in 64 hospitals across ten African countries found that one in two patients died without receiving medical oxygen.[7] However, the global crisis of inadequate oxygen access predates the COVID-19 pandemic. Experts estimate that prior to the pandemic, “nine in ten hospitals in LMICs lacked access to pulse oximetry and oxygen therapy and only 20% of those patients who needed medical oxygen received it”.[8] One pre-pandemic study of Malawi public sector secondary and tertiary referral hospitals found that 23% lacked adequate oxygen access in the general medical wards.[9]

Improving access to medical oxygen is now recognized as a global health priority. In May 2023, the World Health Assembly ratified Resolution 76.3*, Increasing access to medical oxygen*.[3] The Resolution urges WHO Member States to consider a list of 20 actions, including “set up, as appropriate, national and subnational medical oxygen systems in order to secure the uninterrupted provision of medical oxygen to health care facilities at all levels including both rural and urban set-ups” and “assess the scale of medical oxygen access gaps in their health systems, including at subnational- and local-level health facilities.”[3]

In November 2020, with support from Unitaid, the non-profit organization Partners In Health (PIH) launched the Building Reliable Integrated and Next Generation Oxygen Services (BRING O2) project to accelerate access to medical oxygen in Lesotho, Madagascar, Malawi, Peru, and Rwanda.[10] BRING O2’s activities in Lesotho included the planning and implementation of a novel oxygen distribution network, linking multiple oxygen production hubs to recipients throughout the country and leveraging real-time data to align supply with demand.

We conducted a longitudinal analysis of the BRING O2 oxygen distribution network in Lesotho to characterize the operation and impact of a novel oxygen delivery system and better understand oxygen demand dynamics across multiple hospitals.

## METHODS

### Study design and setting

We conducted a longitudinal analysis of the implementation of a national oxygen distribution network in Lesotho from November 2022 through January 2024 by abstracting data from tracking logs used by the oxygen distribution network.

The Kingdom of Lesotho is a small, lower-middle income country of approximately 2.3 million people[11] bordered on all sides by South Africa. Lesotho’s Ministry of Health oversees a network of public health facilities and healthcare services including district and tertiary hospitals as well as Village Health Posts providing community-level services.[12] At the outset of 2020, Lesotho depended entirely on imported liquid oxygen tanks from South Africa. However, during the pandemic, many countries including South Africa halted oxygen exports, exacerbating critical shortages in Lesotho.[13]

In November 2020, the non-profit health organization Partners In Health (PIH) installed Lesotho’s first oxygen production plant at the Botsabelo multidrug resistance-tuberculosis (MDR-TB) Hospital in the capital city, Maseru, and started producing and distributing oxygen to heath facilities throughout the country.[14] In December 2021, with funding from Unitaid, PIH launched the multi-country BRING O2 project to increase access to oxygen.[10] In Lesotho, BRING O2 recruited and trained biomedical engineers and technicians, maintained and repaired oxygen plants, and implemented a national oxygen distribution network. Project goals were aligned with and developed alongside Lesotho’s *National Medical Oxygen Strategy & Scale Up Plan*[15] through participation in the National Medical Oxygen Taskforce, meetings with government officials, and discussions with key stakeholders.

### Oxygen distribution network

The oxygen distribution network was designed to ensure availability of medical oxygen to healthcare facilities in Lesotho lacking sufficient capacity to meet patient demand. To accomplish this objective, PIH provided logistics, technical expertise, and specialized equipment to connect hubs—facilities equipped with oxygen generation plants capable of supplying excess oxygen—to healthcare organizations facing oxygen shortages.

The network operates by retrieving filled oxygen cylinders from hubs (figure 1a) and transporting them to recipients based on identified needs (figure 1b). At the time of delivery, any empty cylinders are collected from recipients and transported back to a hub for refilling (figure 1c). PIH and the BRING O2 project covered the costs of oxygen distribution and did not charge any fees to network participants. Depending on monthly fluctuations in supply and demand, it is possible for a given facility to participate in the network as both a hub and recipient. For example, a hub might transition to a recipient role if its oxygen generation system malfunctions or if onsite oxygen demand exceeds its plant’s production capacity.

**Figure 1:**
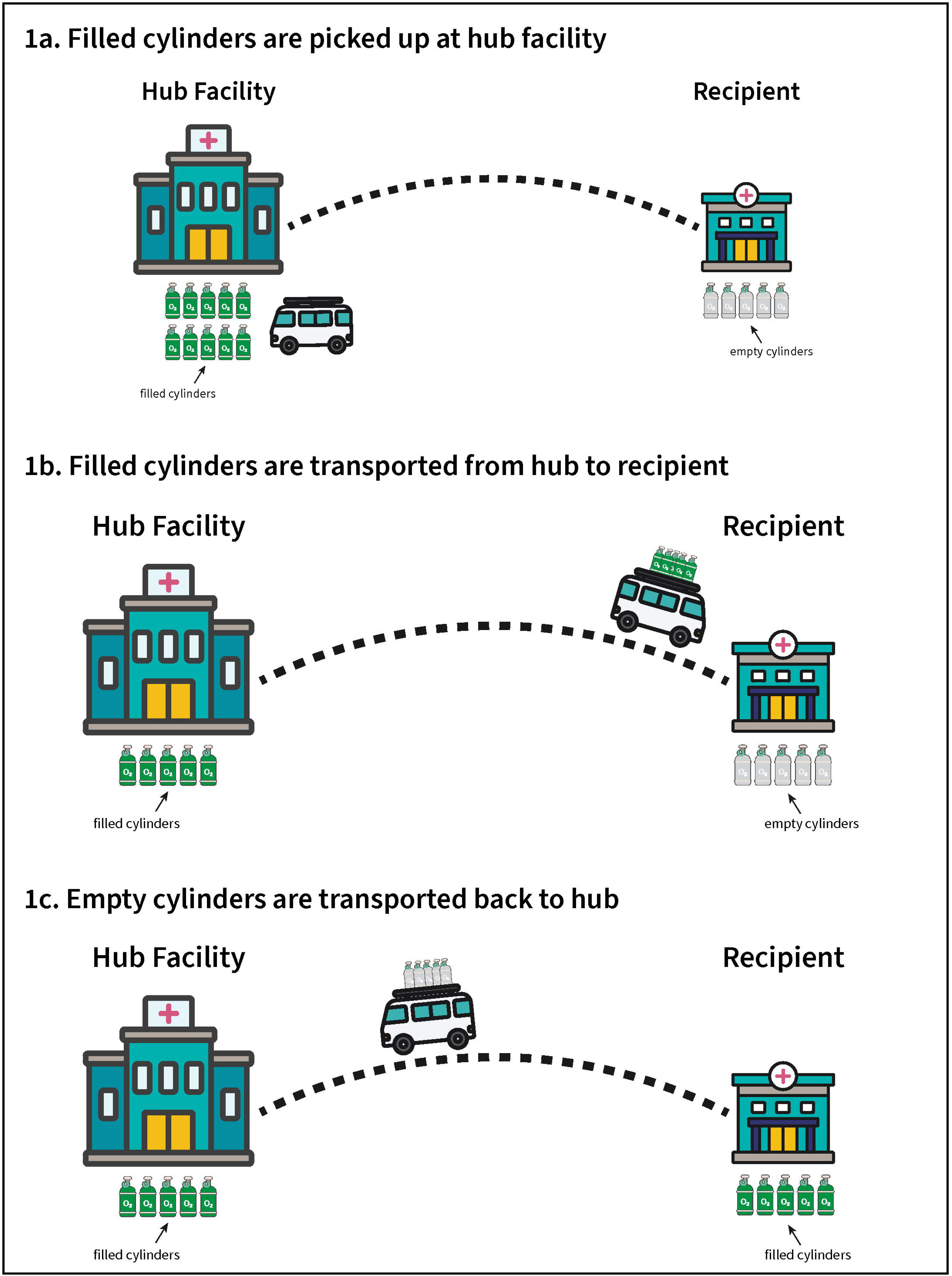
Operation of oxygen distribution network. A) Oxygen cylinders are filled at hubs and retrieved for distribution. B) Filled oxygen cylinders are transported to recipients. C) Filled oxygen cylinders are delivered to recipients and empty cylinders are collected for transport back to a hub for refilling.

Key PIH-Lesotho staff were involved in all stages including planning, oversight, maintenance, and distribution. A driver, trained in safety protocols for handling and transporting oxygen, transported cylinders between oxygen hubs and recipients. Two biomedical engineers were responsible for maintenance of the Botsabelo MDR-TB hospital pressure swing adsorption (PSA) oxygen plant and provided oversight of operations. Six oxygen plant assistant operators filled cylinders at the Botsabelo MDR-TB Hospital oxygen plant and accompanied the driver on delivery runs. A data manager tracked oxygen supply and demand across the system to inform oxygen allocation.

Under the BRING O2 project, PIH procured essential equipment for the network including a flatbed truck that was modified with a rear-mounted hydraulic loading platform and mountable oxygen cylinder racks. Two-row steel chains were installed on all racks to secure and align the oxygen cylinders during transport. These racks also featured a removable small cylinder rack designed to accommodate smaller oxygen cylinders. The truck had a total capacity of 60 cylinders, with 15 single-sided rack positions on the sides of the cargo bed and 30 double-sided cylinder positions in the middle. PIH provided additional equipment such as oxygen cylinders (100 purchased by PIH), trolleys for transporting single cylinders (8 purchased by PIH), oxygen flowmeters, and oxygen regulators.

Implementing the network also required infrastructure upgrades to ensure safe storage and transport of cylinders. PIH constructed a cylinder storage unit adjacent to the Botsabelo MDR-TB hospital oxygen plant. For ease of oxygen cylinder transport, PIH also built concrete loading ramps at the Botsabelo MDR-TB hospital.

Prior to the network launch, PIH performed an initial needs assessment at recipient facilities. Assessment of need was based on demand at the Botsabelo MDR-TB hospital and requests submitted by other facilities. If demand was expected to exceed supply, outreach to other hubs was initiated. During network operation, cylinder counts were performed on a routine basis. Specific protocols were established for each hub facility: at Mafeteng, obtaining a memo from the Ministry of Health in advance was necessary for cylinder pick up, while at Paray and Motebang discussions were held with administrators prior to cylinder pick-up and delivery. The network was originally designed and launched with 8 recipients. Additional recipients were added over the course of the study period based on unsolicited expressions of interest to PIH.

Tracking logs captured the date, filling hub, recipient, and number and size of cylinders for each delivery. Cylinder sizes were denoted by volume of water displaced (e.g., 50-litre, 40-litre, etc.). These logs were regularly updated and reviewed periodically for accuracy by the PIH team. Any discrepancies were reviewed and corrected by BRING O2 staff. We report data recorded from the tracking logs starting from the launch of the network in November 2022 through January 2024. Any facility with missing tracking log data was coded as receiving no cylinder deliveries for the given month.

### Variables

We recorded the quantity of cylinders delivered by hub and recipient classified by size and associated oxygen volume, expressed in cubic meters (m^3^) at atmospheric conditions. Oxygen volume was calculated from cylinder count data, assuming a filled cylinder pressure of 14,478.996 KPa and atmospheric pressure of 101.325 KPa, utilizing the following formula:

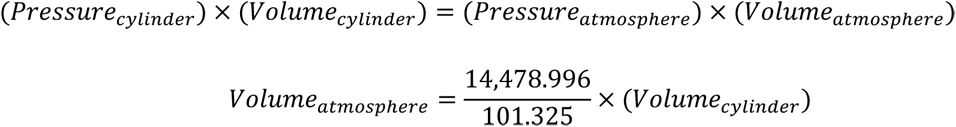

Distance between hubs and recipients were obtained by inputting location information (latitude and longitude) into Google Maps.[16]

### Statistical analysis

Study data were abstracted from network tracking logs and analysed in Stata (Release 16). Continuous and ordinal variables were summarized by medians and ranges. Categorical variables were described using frequencies and proportions. In subgroup analyses, we included only network recipients with inpatient beds.

### Study reporting

The research presented in this manuscript adheres to the reporting standards of the Strengthening the Reporting of Observational Studies in Epidemiology (STROBE) Statement guidelines for reporting observational studies.[17]

### Role of funding source

The funders had no role in study design; collection, analysis, and interpretation of data; decision to publish; or preparation of the manuscript.

### Patient and public involvement

Due to the nature of the study, patients and public involvement in the design, conduct, reporting or dissemination plans of our research was not applicable.

### Author reflexivity statement

The authors of this paper have submitted a Reflexivity Statement (supplemental appendix A). This statement allows researchers to describe how their work engages with researchers, communities, and environments in the country of study.

## RESULTS

The oxygen distribution network included four hub facilities with on-site oxygen production plants where empty cylinders were filled with oxygen and distributed to off-site recipients. Oxygen was delivered to a total of 21 unique recipients, including two of the four hub facilities (Botsabelo MDR-TB Hospital and Motebang Hospital) which received cylinders during periods of oxygen plant down-time and excess on-site demand. The network had broad geographical coverage, with recipients in nine of Lesotho’s 10 districts (figure 2). The number of unique recipients receiving cylinders through the network increased from 6 in the fourth quarter (October to December) of 2022 to 12 in the fourth quarter of 2023, with a maximum of 14 recipients the third quarter (July to September) of 2023 (figure S1, supplemental appendix B).

**Figure 2:**
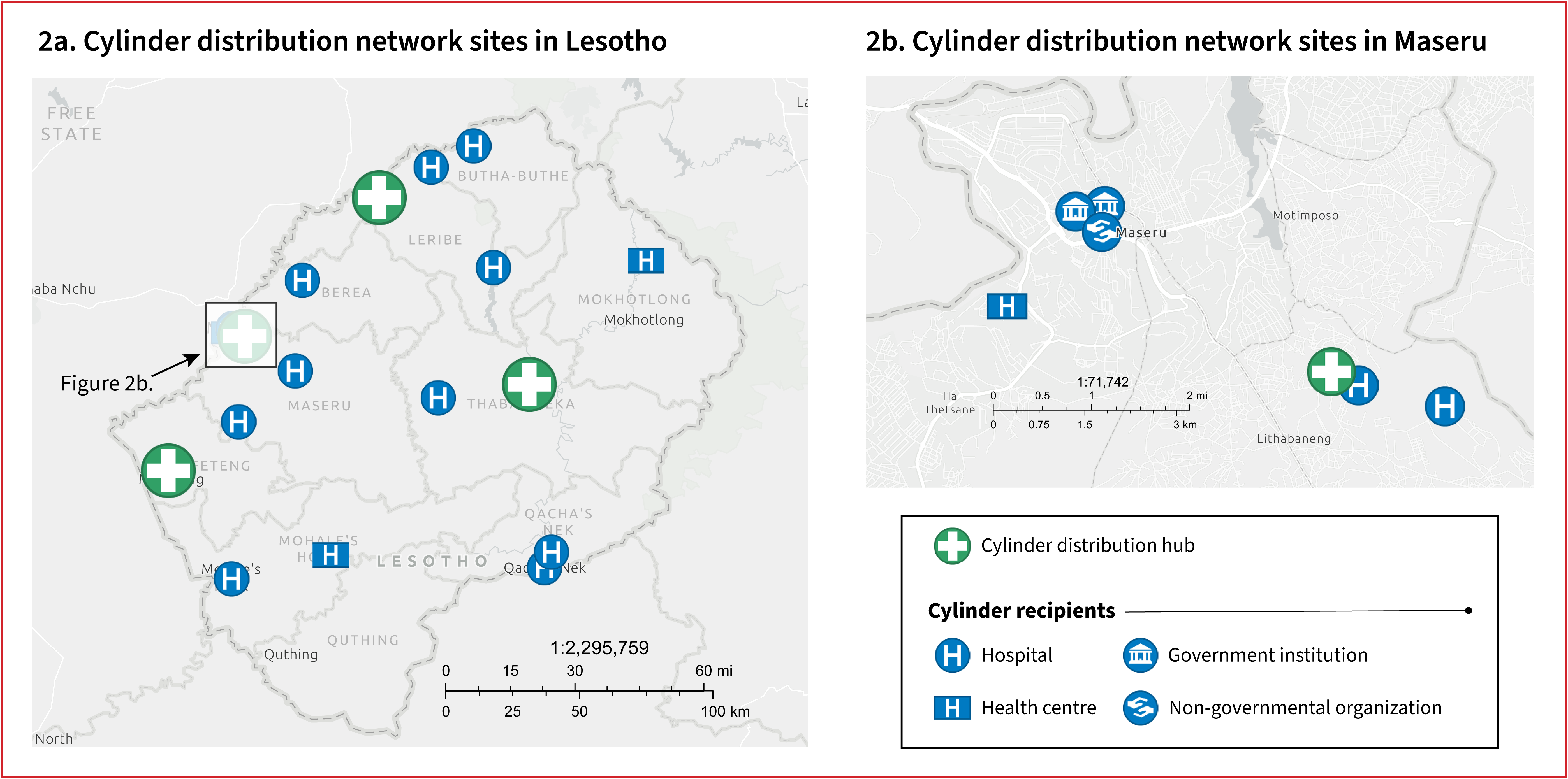
Map of cylinder distribution sites in Lesotho and Maseru. A) Country map of Lesotho showing locations of cylinder network hubs and recipients B) City map of Maseru showing locations of cylinder network hubs and recipients To protect individual privacy, the location of one recipient (home oxygen delivery) is not included

The network distributed a total of 1,565 filled cylinders containing 9,619.23 cubic meters (m^3^) oxygen over the 15-month study period (table 1). Botsabelo MDR-TB Hospital served as the primary hub, providing 1,020 (65.2%) cylinders containing a total of 6,059.82 m^3^ oxygen (63.0%). The other three hubs supplemented capacity during periods of high demand or when the Botsabelo oxygen plant experienced down-time. The median distance by road from hubs to recipients was 87.7 kilometres (range: 0.5 to 328.0). The network relied primarily on large capacity cylinders, with 50- and 40-litre sizes accounting for 60.1% and 30.5% of all distributed cylinders, respectively.

**Table 1.**
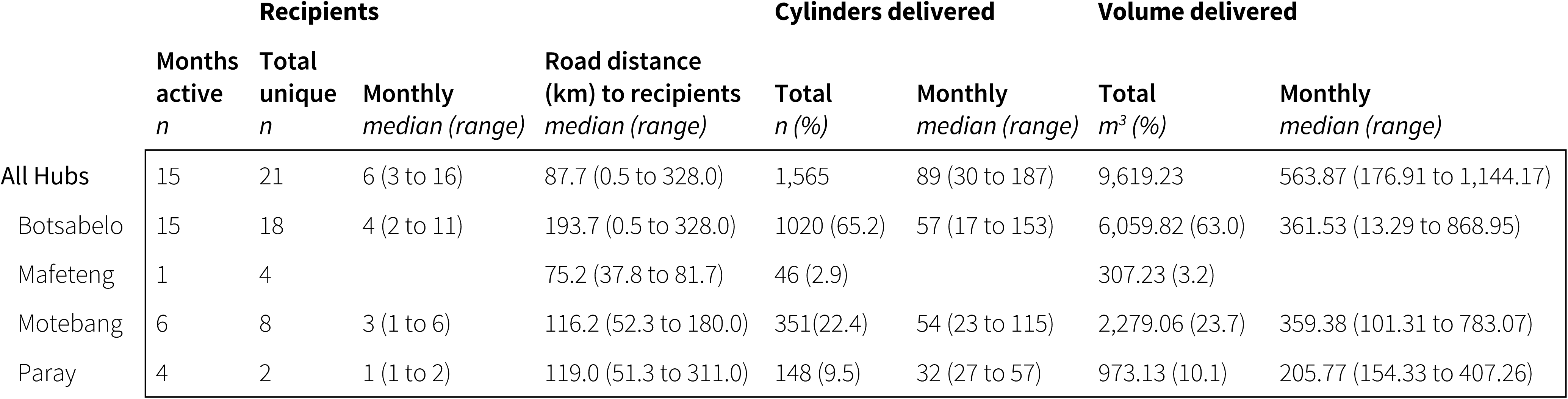
Cylinder network hub facilities.

### Oxygen distribution by hub and recipient

Figure 3a shows the number of oxygen cylinders distributed by month and hub. The Botsabelo MDR-TB Hospital oxygen plant functioned as the primary hub with supplemental contributions from oxygen plants at Motebang Hospital, Mafeteng Hospital, and Paray Hospital. While 13 (61.9%) recipients received all oxygen deliveries from a single network hub, 8 (38.1%) recipients received oxygen deliveries from multiple hubs during the study period (Figure 3b). Botsabelo MDR-TB Hospital served as a hub for all recipients except Nts’ekhe Hospital.

**Figure 3:**
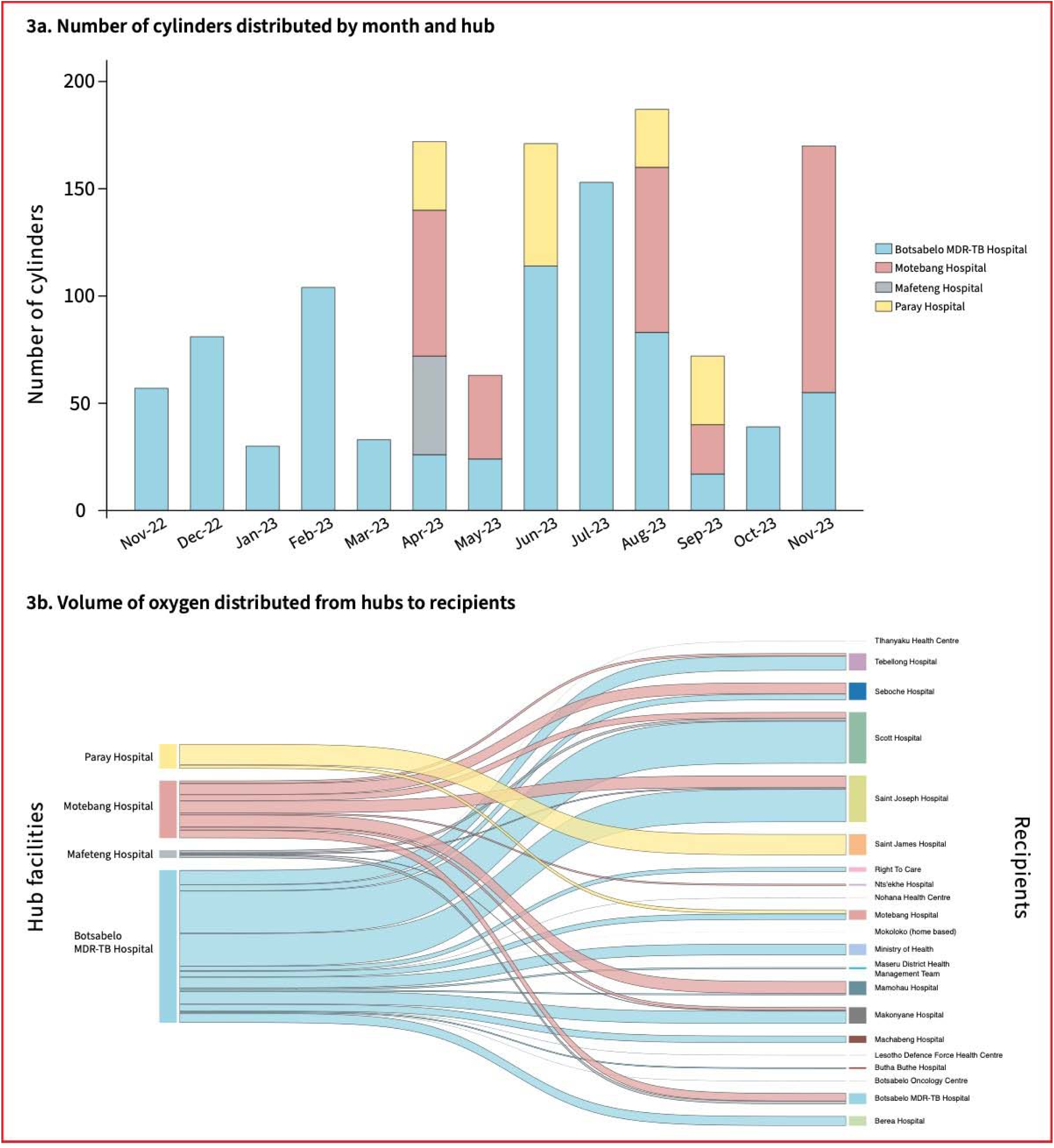
Oxygen distribution by hubs and recipients over time. A) Bar chart demonstrating number of cylinders distrubted per month broken down by hub B) Sankey diagram demonstrating movement of oxygen volume (m3) from hubs to recipient facilities during the study period. Line thickness represents relative volume, with thicker lines indicating more volume.

Figures 3a and 3b demonstrate how integrating multiple hubs into a single network of recipients made it possible to compensate for periods when oxygen plants break down. For roughly two months starting in June 2023, the Motebang oxygen plant broke down and was no longer able to produce oxygen. During this period, Motebang Hospital was unable to fill cylinders for the network and also faced an oxygen shortage for its patients. In response, the Botsabelo and Paray hubs increased production to meet network demand, including the delivery of over 40 cylinders to sustain Motebang Hospital’s oxygen supply until the oxygen plants were repaired.

### Volume per hospital bed

There was wide variation in the amount of oxygen delivered to recipients. For the 13 recipient facilities with inpatient beds, the median monthly volume of oxygen delivered per hospital bed was 1.43 m^3^ (IQR: 0.57 to 2.31) during the study period (table 2). Scott Hospital received the largest number of cylinders and associated oxygen volume among all recipients (332 cylinders and 2038.99 m^3^, respectively), while Saint James Hospital had the highest median monthly volume of oxygen delivered per hospital bed among non-hub facilities at 2.06 m^3^ (IQR: 1.54 to 2.64).

**Table 2.** Cylinder network recipients.

|  | Months<br>active<br>*n* | Inpatient<br>beds<br>*n* | Cylinders received |  | Volume received |  | Monthly per<br>inpatient bed<br>median (range) |
| --- | --- | --- | --- | --- | --- | --- | --- |
|  |  |  | Total | Monthly<br>median (range) | Total<br>*m*<sup>3</sup> | Monthly<br>median (range) |  |
|  |  |  | *n* |  |  |  |  |
| Hospitals |  |  |  |  |  |  |  |
| Berea Hospital | 5 | 128 | 69 | 9 (4 to 37) | 380.10 | 51.44 (6.43 to 199.34) | 0.40 (0.05 to 1.56) |
| Botsabelo MDR-TB Hospital | 3 | 24 | 63 | 16 (8 to 39) | 430.12 | 114.32 (57.16 to 258.64) | 4.76 (2.38 to 10.78) |
| Butha Buthe Hospital | 1 | 129 | 16 |  | 55.73 |  |  |
| Machabeng Hospital | 4 | 96 | 42 | 8 (6 to 20) | 284.79 | 50.94 (40.01 to 142.90) | 0.53 (0.42 to 1.49) |
| Makonyane Hospital | 10 | 37 | 107 | 8 (1 to 26) | 651.18 | 52.87 (1.29 to 172.90) | 1.43 (0.04 to 4.67) |
| Mamohau Hospital | 7 | 51 | 101 | 20 (13 to 30) | 551.87 | 110.32 (46.44 to 200.06) | 1.97 (0.83 to 7.15) |
| Motebang Hospital | 3 | 192 | 53 | 27 (12 to 41) | 378.68 | 189.34 (85.74 to 292.94) | 0.60 (0.45 to 0.76) |
| Nts'ekhe Hospital | 1 | 134 | 10 |  | 71.45 |  |  |
| Saint James Hospital | 4 | 100 | 128 | 32 (27 to 37) | 830.23 | 205.77 (154.33 to 264.36) | 2.06 (1.54 to 2.64) |
| Saint Joseph Hospital | 13 | 120 | 267 | 24 (7 to 45) | 1,841.94 | 189.34 (50.01 to 305.80) | 1.25 (0.42 to 2.29) |
| Scott Hospital | 14 | 102 | 332 | 30 (8 to 67) | 2038.99 | 182.91 (3.43 to 448.70) | 1.15 (0.03 to 4.05) |
| Seboche Hospital | 7 | 88 | 125 | 20 (13 to 28) | 691.05 | 102.89 (62.16 to 188.62) | 1.17 (0.48 to 2.14) |
| Tebellong Hospital | 10 | 50 | 107 | 9 (3 to 23) | 682.47 | 64.30 (7.72 to 135.75) | 1.29 (0.15 to 2.72) |
| Health Centres/Clinics |  |  |  |  |  |  |  |
| Botsabelo Oncology Centre | 1 |  | 1 |  | 7.14 |  |  |
| LDF Health Centre | 2 |  | 7 | 4 (1 to 6) | 9.00 | 4.50 (1.29 to 7.72) |  |
| Nohana Health Centre | 2 |  | 7 |  | 9.00 |  |  |
| Tlhanyaku Health Centre | 2 |  | 4 |  | 5.14 |  |  |
| Organizations/Other |  |  |  |  |  |  |  |
| Maseru DHMT | 5 |  | 22 | 4 (1 to 10) | 73.31 | 3.00 (2.57 to 58.02) |  |
| Ministry of Health | 3 |  | 71 | 30 (1 to 40) | 442.84 | 214.34 (1.29 to 227.21) |  |
| Right To Care | 3 |  | 32 | 14 (1 to 17) | 182.91 | 80.02 (5.72 to 97.17) |  |
| Home Care* | 1 |  | 1 |  | 1.29 |  |  |

MDR-TB: Multidrug resistant tuberculosis
LDF: Lesotho Defence Force
DHMT: District Health Management Team
\*Direct delivery to patient home

The Paray Hospital and Mafeteng Hospital hubs did not receive any oxygen deliveries during the study period, while Motebang Hospital had a relatively low demand per bed of 0.60 m^3^ (IQR: 0.45 to 0.76) during the 3 months it received deliveries. Over the 3 months that Botsabelo MDR-TB Hospital received outside cylinder deliveries, the demand per bed was relatively high 4.76 m^3^ (IQR: 2.38 to 10.78) compared to other facilities. Figure S2 (supplemental appendix B) illustrates the increasing trend over time in oxygen demand by bed at the 13 recipients with inpatient facilities.

## DISCUSSION

This longitudinal analysis of an oxygen distribution network established under PIH’s BRING O2 project, demonstrates the feasibility and effectiveness of a novel approach to ensuring oxygen access within an LMIC. By sourcing oxygen from multiple hubs, the network provided a resilient oxygen supply to 21 recipients located across nine of Lesotho’s ten districts. Over 15 months, the network provided 9,619.23 m^3^ of oxygen, equivalent to 3,036 patient days of oxygen therapy (assuming a mean oxygen flow rate of 2.2 litres/min).[18] This study establishes a proof-of-concept for oxygen distribution networks as a key tool for achieving oxygen security within an LMIC.

Furthermore, by integrating multiple hubs, the network model may increase efficiency by reducing required travel distance for oxygen delivery to peripheral sites. A prior study of single-hub networks reported transporting oxygen to recipients located up to 200 km away from the hub.[19] In comparison, the Lesotho oxygen distribution network Lesotho distributed oxygen a median distance of 87.7 km.

Over the study period, increasing numbers of healthcare facilities requested to join the network and a trend toward increasing oxygen demand per bed over time was observed. Similarly, a prior study found that distribution systems increased oxygen procurement at recipient facilities by 112 to 220%.[19] Together, these findings suggest that making oxygen available through distribution networks has to the potential to unlock significant additional demand.

Implementing the oxygen distribution network required investments in biomedical staff, equipment, oxygen production and storage infrastructure, and monitoring systems. While multiple studies have evaluated multi-dimensional interventions to improve oxygen access at the facility level,[6,20,21] there is limited evidence from regional or national level interventions. This study adds to the body of evidence supporting interventions that take a multi-dimensional approach to increasing oxygen access.

Ensuring continuous oxygen access is essential; even brief shortages can result in significant morbidity and mortality. However, national oxygen strategies do not always account for the limited fungibility of oxygen when aggregating oxygen supply and demand at the national level.[22–24] A more granular approach, accounting regional and facility-level differences in oxygen sources and delivery is required to prevent oxygen shortages. By operating at a national level while also monitoring real-time feedback from individual hubs and recipients, the network was able to adjust deliveries to align supply and demand, providing a mechanism for ensuring oxygen security.

Dynamic sourcing across multiple hubs enabled the Lesotho oxygen distribution network to provide an uninterrupted supply of oxygen to recipients despite fluctuations in supply and demand. When the Motebang oxygen plant broke down in June – August 2023 halting its oxygen production, the Botsabelo and Paray hubs responded by increasing production, sending over 40 cylinders to sustain Motebang Hospital’s oxygen supply until the plant was repaired. One prior study[19] and several national oxygen strategies[15,22,24] have discussed implementing “hub-and-spoke” oxygen cylinder distribution systems. These systems create cylinder delivery links (i.e., spokes) to recipients from a single oxygen production source (i.e., hub). However, this model relies on a single oxygen production source (e.g., a PSA plant), making the entire system highly vulnerable to fluctuations in supply and demand arising from common events including power failures, plant breakdowns, and viral respiratory illness outbreaks. By employing a network model with multiple hubs, this study provides a proof-of-concept mechanism for increasing the resiliency and security of medical oxygen supply at the regional and national level. Additionally, this system could serve as an early warning mechanism for mismatches in oxygen supply and demand, particularly in dynamic situations such as a pandemic respiratory illness. These findings have the potential to inform future oxygen ecosystem strengthening efforts by policymakers, funders, and implementers.

Our study also noted variability in oxygen demand in Lesotho across facilities and over time. This variation may be due to several factors, including oxygen sourcing (i.e., reliance on network deliveries versus alternate sources such as oxygen concentrators), hospital infrastructure (e.g., installation of oxygen piping), and clinical practice patterns. Indeed, one study reported a 53% increase in oxygen demand following the provision of clinical training.[19]

This work also provides insights for a potential approach to generating real-world estimates of regional and national oxygen demand. Over the study period, we calculated a median oxygen demand by volume of 1.43 m^3^ per month per hospital bed, or approximately 2,915.77 m^3^ per month in total national oxygen demand for Lesotho’s 2,039 hospital beds. However, this is likely an underestimate as the network was not the sole oxygen supplier to the identified hospital facilities during the study period. This estimate differs from that of Lesotho’s national strategy, which estimated a total oxygen demand of 19.3 m^3^ per hospital bed or 39,375 m^3^ monthly oxygen need across all hospitals.[15] Additionally, our calculations do not account for patients with hypoxemia that are not identified at facilities or are unable to engage with the healthcare system. Future studies should prioritize collecting facility-level data on all oxygen sources and patient-level data on hypoxemia prevalence. This information can be combined with oxygen delivery data to provide a more nuanced understanding of oxygen demand patterns and inform targeted strategies for improving patient care and strengthening oxygen ecosystems.

This work opens several additional avenues for implementation and investigation. The multi-hub approach used by the Lesotho network could be expanded to include both PSA plants and liquid oxygen (LOX) sources as hubs within a single integrated network, providing increased redundancy and flexibility. These activities could be complemented through concurrent implementation of proven facility-level interventions.[6,25] In Lesotho, next steps include strengthening complementary elements of its oxygen ecosystem, such as enhancing hospital infrastructure to ensure more efficient and reliable oxygen delivery, which may involve installing oxygen piping and upgrading essential equipment such as flowmeters and regulators. This study also raises the possibility of leveraging oxygen distribution networks as a platform for data collection and monitoring systems to track and predict oxygen usage at regional and national levels, providing much-needed information to guide efforts by policymakers and oxygen suppliers to ensure oxygen security on a national level.

Several limitations should be acknowledged. Oxygen network services were provided free of charge by PIH and this study did not collect detailed cost data. The lack of cost data may limit the replicability of this intervention. However, a prior study of single-hub oxygen networks across three countries found an estimated cost of US$7.34 per patient treated, though inconsistent revenue was noted as a major challenge.[19] The study authors also suggested that the costs of oxygen distribution could ultimately be passed down onto patients.[19] However, this approach risks placing a financial burden on patients, creating potential misalignment with the principles of Universal Health Coverage.[26] Future work is needed to identify sustainable models that will not place a financial burden on patients.

Another limitation is the use of monthly oxygen delivery as a proxy for oxygen demand. While this approach has been previously employed to estimate oxygen consumption,[19] it is possible that not all delivered oxygen was used for patient care, leading to an overestimate of demand. Alternatively, the fact that recipients were able to obtain oxygen from sources external to the network may lead to underestimating true demand.

## CONCLUSION

The oxygen distribution network established in Lesotho represents a significant advance in oxygen security in resource-limited settings. Using data monitoring systems and inbuilt redundancies, the network was able to align oxygen supply and demand, demonstrating resiliency during the outage of a production hub. In doing so, the network was also able to gain insights into the dynamics of oxygen demand over time. Future efforts should focus on enhancing data collection, characterizing oxygen usage, and strengthening infrastructure to promote sustainable oxygen security.

## Supporting information

Reflexivity statement

## DATA AVAILABILITY STATEMENT

All data collected during the study will be available Immediately following publication, with no end date, to researchers who provide a methodologically sound proposal. Proposals should be directed to; to gain access, data requestors will need to sign a data access agreement.

## ETHICS STATEMENTS

### Patient consent for publication

Not applicable.

## ACKNOWLEDGEMENTS

We extend our deepest gratitude to Lesotho’s Ministry of Health for partnership and guidance throughout the planning and implementation of the oxygen distribution network project and to Unitaid for supporting the BRING O2 project. We also thank all partners in Lesotho who work tirelessly to improve oxygen access for patients.

A subset of data from this study were recently presented in abstract form at the 15th Annual Consortium of Universities for Global Health Conference (March 7-10, 2024; Los Angeles, CA).

## CONTRIBUTORS

MN, ATA, and PDS conceived and designed the study. MN, ATA, EG, JM, and PDS acquired funding. MN, ATA, JP, JM, and PDS were responsible for project supervision. PT, NS, MP, WH, and EG administered the project. PT, MT, FS, TM, and TC collected and curated the data. MN, ATA, PT, TM, MT, JP, and PDS have accessed and verified the data. MN, ATA, JL, and PDS did the analyses and wrote the first draft. All authors reviewed and edited the manuscript. All authors approved the final manuscript version. MN and PDS had final responsibility for the decision to submit for publication.

## FUNDING

This work was supported by Unitaid (grant SPHQ15-LOA-045). The funders had no role in study design; collection, analysis, and interpretation of data; decision to publish; or preparation of the manuscript.

## COMPETING INTERESTS

The BRING O2 project was supported by Unitaid, grant number SPHQ15-LOA-045, for oxygen ecosystem strengthening in five countries including Lesotho. MN, PT, MT, FS, TM, NS, MP, JP, WH, EG, and PDS all report support from this grant. PDS reports grants or contracts from the University of California-San Francisco/Sustaining Technical and Analytic Resources, Wyss Foundation, Clinton Health Access Initiative, FHI360, The World Bank Group, and the World Health Organization; consulting fees from the University of California-San Francisco; unpaid advisory roles in the Global Oxygen Alliance and the World Health Organization Expert Working Group on Technical Specifications for Medical Oxygen Systems; and equity in Nininger Medical.

## Notes

### Summary of Updates

Figure 3 has been resized to fit page.

