## Supplementary material for "Implementation of a national oxygen distribution network in Lesotho: a longitudinal analysis": Reflexivity statement

1. How does this study address local research and policy priorities?

This project was specifically developed to increase oxygen access via tailored solutions in the Lesotho context. BRING O2 included the Lesotho Ministry of Health and other key stakeholders the planning and implementation of project activities. The Lesotho Ministry of Health approved the BRING O2 project, reflecting the prioritization of oxygen availability in Lesotho.

2. How were local researchers involved in study design?

Colleagues from Bo-Mphato Litšebeletsong Tsa Bophelo in Lesotho and the Partners In Health (PIH) coordination site in the United States collaborated in the design and implementation of the oxygen cylinder network and the design of this study. This included a series of in-person meetings held in Maseru, Lesotho which included team members from Bo-Mphato Litšebeletsong Tsa Bophelo and the PIH coordination site and ongoing open communication across sites. Local researchers (MN, AA, PT, MT, FS, TM, NS, MP) were key members of the study team, contributing expertise running domestic and international research collaborations, knowledge of Lesotho’s oxygen environment, and/or experience providing direct clinical care.

3. How has funding been used to support the local research team?

BRING O2 provided direct salary support for all key staff on this project and provided funds for the purchase of new equipment to prevent inappropriate reallocation of existing equipment.

4. How are research staff who conducted data collection acknowledged?

All research staff who participated in data collection at the healthcare sits were included as authors. Please see acknowledgements for further descriptions of individual roles.

5. Do all members of the research partnership have access to study data?

Unrestricted full access to the database was granted to colleagues from Bo-Mphato Litšebeletsong Tsa Bophelo and the PIH coordination site.

6. How was data used to develop analytical skills within the partnership?

The data analysis plan was initially outlined by colleagues from Bo-Mphato Litšebeletsong Tsa Bophelo and further refined through discussions with PIH coordination site colleagues. Results were shared with Bo-Mphato Litšebeletsong Tsa Bophelo. Discussions (in-person, virtual, and email) were held at multiple points during the study to provide Bo-Mphato Litšebeletsong Tsa Bophelo colleagues with insight into the analysis process, allowing them to provide feedback and offer interpretations. All collaborators had unrestricted access to the study database.

7. How have research partners collaborated in interpreting study data?

Colleagues from Bo-Mphato Litšebeletsong Tsa Bophelo, the PIH coordination site, and Brigham and Women's Hospital/Harvard Medical School (BWH/HMS) in the United States collaborated in data analysis and interpretation. Specifically, the study database was designed and developed during a series of in-person meetings held in Maseru, Lesotho between colleagues from Bo-Mphato Litšebeletsong Tsa Bophelo and the PIH coordination site.

The data analysis plan was initially outlined by colleagues from Bo-Mphato Litšebeletsong Tsa Bophelo and shared with colleagues from the PIH coordination site. The data analysis plan was further refined during a series of discussions and emails among these colleagues. Colleagues from the PIH coordination site and BWH/HMS conducted analyses according to the analysis plan and shared all outputs with colleagues from Bo-Mphato Litšebeletsong Tsa Bophelo. Data interpretation was conducted through a series of discussions (in-person, virtual, and email) among colleagues from Bo-Mphato Litšebeletsong Tsa Bophelo, the PIH coordination site and BWH/HMS.

8. How were research partners supported to develop writing skills?

Colleagues from Bo-Mphato Litšebeletsong Tsa Bophelo, the PIH coordination site and BWH/HMS collaborated in manuscript preparation. Specifically, the first, second, and senior authors collaborated to develop the initial draft of the manuscript. Mentorship on manuscript preparation was provided by senior authors. All co-authors reviewed and edited the manuscript for key intellectual content, in particular elaborating on presentation of context and on policy implications and recommendations.

9. How will research products be shared to address local needs?

The findings of this study, the broader BRING O2 project, have been shared with a variety of stakeholders during meetings and discussions. Additionally, a written layperson summary was developed and distributed to key partners in Lesotho, including staff at the Lesotho Ministry of Health. When the manuscript is published, the paper copy and electronic link will be shared with the Ministry of Health and implementing partners.

10. How is the leadership, contribution and ownership of this work by LMIC researchers recognised within the authorship?

The authorship team was composed of researchers from both LMICs and HICs. MN and ATA are recognized as first and second authors, respectively, for their work conceiving, designing, supervising, and administering the project with assistance from the project’s senior authors.

11. How have early career researchers across the partnership been included within the authorship team?

The authorship team included early career researchers (AA, JL, and TC) who were equal partners in the project. These individuals contributed to the project creation and planning, data collection, manuscript drafting and review.

12. How has gender balance been addressed within the authorship?

Eight authors are male (MN, AA, MT, FS, TM, NS, WH, and PS) and seven authors are female (PT, JL, MP, JP, EG, TC, and JM).

13. How has the project contributed to training of LMIC researchers?

The authorship team is comprised of both senior and junior researchers in the United States and Lesotho. Training, mentorship, and development of long-term skills were specifically included within BRING O2 activities.

Across five implementing countries, BRING O2 trained over 600 healthcare workers. The first author provided longitudinal one-on-one mentorship to colleagues at Bo-Mphato Litšebeletsong Tsa Bophelo on data collection, management, and analysis. Implementing this research project additionally offered practical experience in the grant and research execution.

14. How has the project contributed to improvements in local infrastructure?

The BRING O2 project built multiple structures to support oxygen infrastructure, including cylinder storage areas and cement loading ramps. All construction was planned and implemented with close coordination and collaboration with key stakeholders.

15. What safeguarding procedures were used to protect local study participants and researchers?

Primary patient data was not collected as part of this project; therefore, this question is less directly applicable. However, all construction was planned and implemented with close coordination and collaboration with key stakeholders.
