## Supplemental figures for "Implementation of a national oxygen distribution network in Lesotho: a longitudinal analysis"

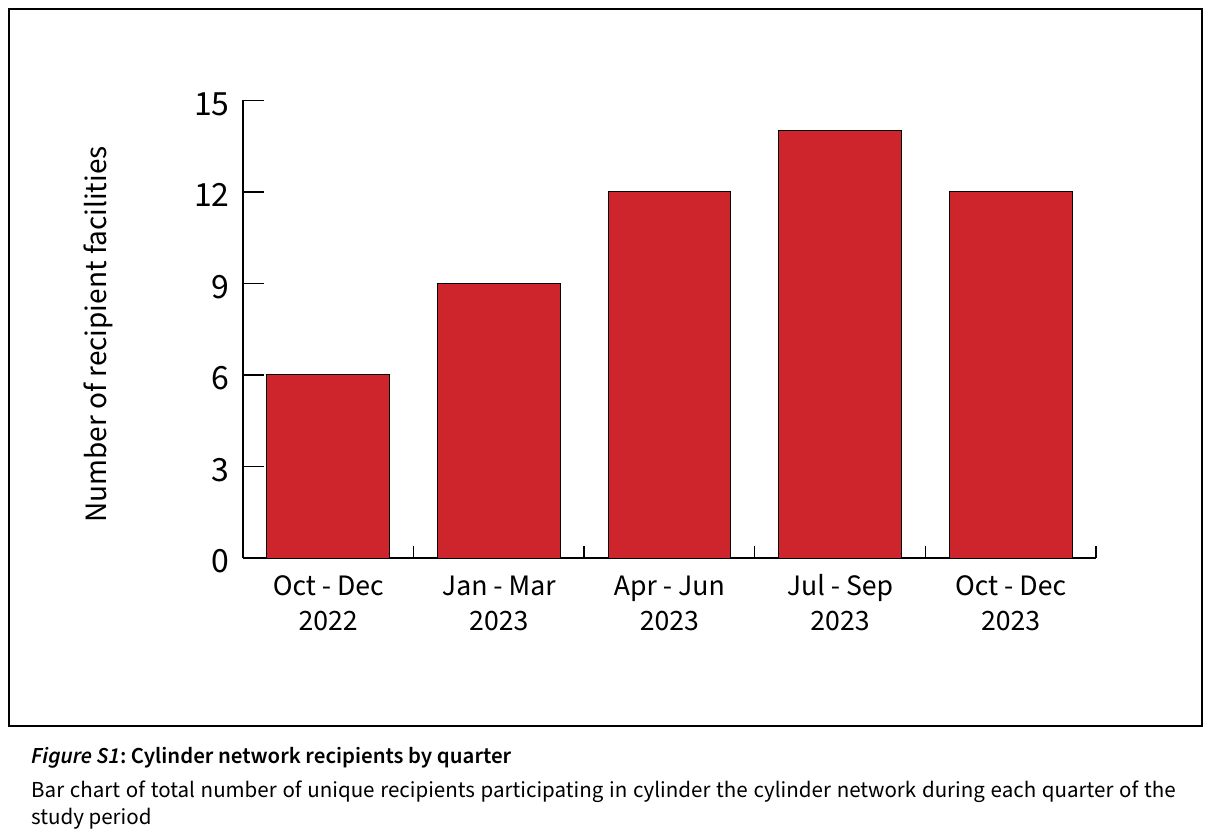

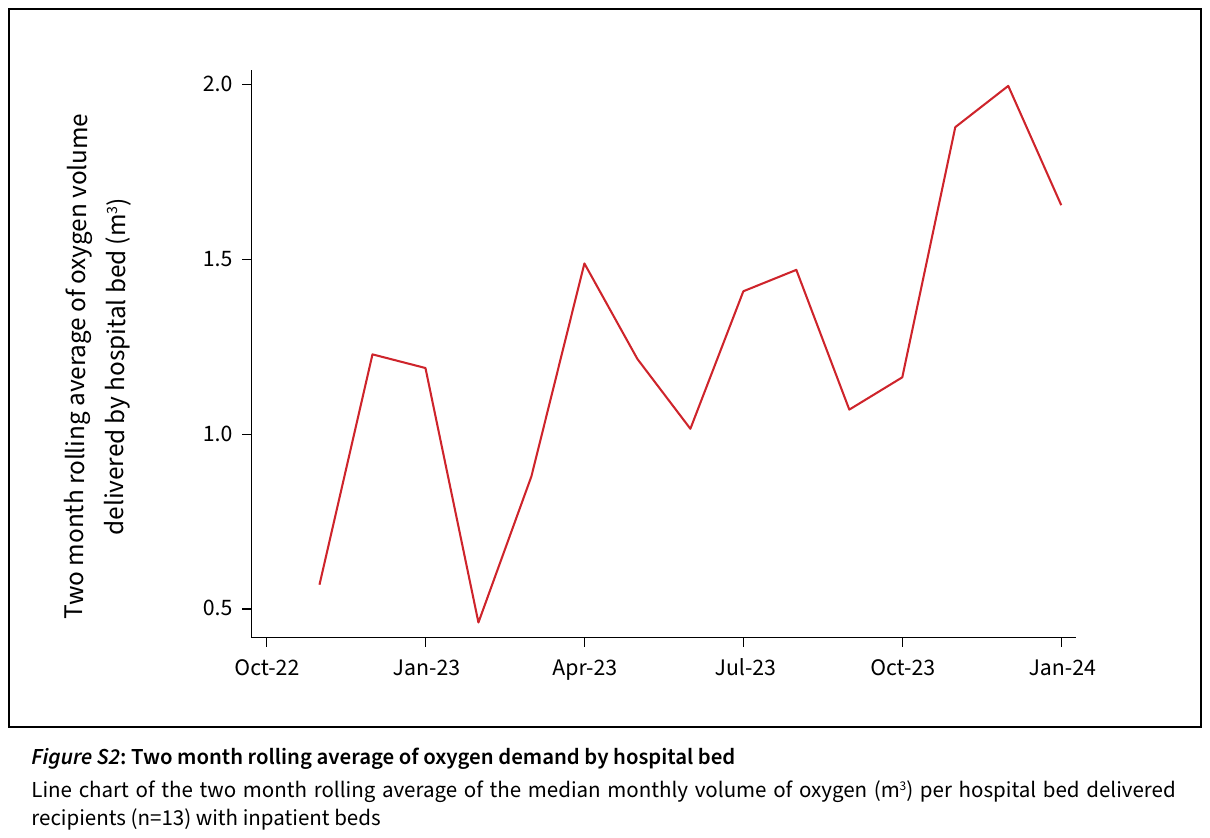
